# Social Adversity, Systemic Inflammation, and the Ticking of the Biological Aging Clocks in Men and Women

**DOI:** 10.64898/2026.07.20.26358488

**Authors:** Cesar Higgins Tejera, Rose Noroozi, Keenan A. Walker, Leah H. Rubin, Kathryn C. Fitzgerald

## Abstract

**Objectives:** We tested how multi-level socioeconomic disadvantage relates to biological aging and systemic inflammation in women and men from the population-based Canadian Longitudinal Study on Aging (CLSA).

**Methods:** We examined cross-sectional data from 8,516 CLSA participants with baseline measures on systemic inflammatory biomarkers (C-reactive protein, interleukin-6, and tumoral necrosis factor-α) and biological aging (metabolomic and six DNA methylation [DNAm] age estimates). Plasma samples underwent metabolomic profiling by Metabolon, Inc. Metabolomic age was estimated separately in males and females using sex-stratified models based on age-correlated metabolite levels. DNAm data generated using the Illumina Infinium MethylationEPIC v1.0 array were used to estimate DNAm age across six established models, including Horvath, Hannum, PhenoAge, GrimAge, GrimAge2, and DunedinPACE. We used log-transformed metabolite levels to calculate metabolomic age by sex. We linked education, income, material and social deprivation to biomarkers of systemic inflammation and biological aging stratified by sex using generalized linear models. Multivariable models were adjusted by age, major behavioral risk factors, and chronic conditions.

**Results:** Participants were aged on average of 62.6 years of age, and approximately 50% were females. In multivariable linear adjusted models, we found that in comparison to those earning ≥$100K a year, women earning less <$20K were on average 1.14 (95%CI: 0.46, 1.82) year older with respect to metabolomic age; those earning ≥$20K & <$50K were on average 0.90 (95%CI: 0.26, 1.53) years older; and those earning ≥$50K & <$100K were on average 0.70 (95%CI: 0.05, 1.34) years older. We did not observe this dose response among men. A similar dose-response association was observed for interleukin-6 in both men and women.

**Discussion:** These findings suggest that socioeconomic adversity influences not only inflammatory pathways but also distinct biological aging processes, including metabolomic aging.

## Introduction

Socioeconomic disadvantage manifests biologically in multiple ways: lower income is associated with heightened systemic inflammation and increased cardiometabolic risk, while lower educational attainment is linked to accelerated DNA methylation (DNAm) age and increased dementia risk in older adults.^1–7^ Beyond these individual-level indicators, neighborhood characteristics also exert a significant influence on health. A growing body of evidence indicates that neighborhood environments shape vulnerability to cardiovascular disease, stroke, and dementia.^8–11^ Collectively, these findings underscore that social adversity, encompassing low income, education, and neighborhood-level marginalization and deprivation, are biologically embedded through multiple molecular pathways that contribute to physiological fragility, age-related disease susceptibility, and multimorbidity. Furthermore, women continue to earn substantially less than men, even when employed in comparable positions.^12^ Despite major advances in educational attainment, women remain underrepresented in high-paying occupations and leadership roles. These persistent structural inequities may contribute to observed differences in morbidity and mortality between men and women, potentially reflecting both socioeconomic disadvantage and differential biological responses to chronic stress and adversity.

The advent of high-throughput omics technologies has enabled the quantification of epigenetic and metabolic signatures that estimate an individual’s biological age, as a measure of physiological functioning at the cellular or organ level relative to what is expected for their chronological age.^13–16^ Biological aging and systemic inflammation represent key pathways through which social adversity may exert long-term physiological effects on health. However, the physiological response to social adversity may differ between men and women due to both sex-specific physiological responses to stress and assigned gender roles.^17,18^ Documenting these differences may help clarify socially patterned responses in biological aging and inflammation and improve our understanding of sex dimorphisms in the physiological response to social adversity and their repercussions to multimorbid chronic conditions.

Intriguingly, despite women’s disproportionate exposure to social and economic adversity, several studies report that women exhibit slower biological aging trajectories and reduced immunosenescence compared to men, consistent with their longer life expectancy and more robust immune responses across the lifespan.^19–22^ However, the biological mechanisms underlying these observed differences remain poorly understood. In the present study, we examine how education, income, and material and social deprivation relate to (1) metabolomic and epigenetic aging clocks, as indicators of biological aging, and (2) to systemic inflammatory markers (C-reactive protein, interleukin-6, and tumor necrosis factor-α) in women and men from a large, population-based study. Our goal was to characterize specific pathways through which social adversity becomes biologically embedded and influences underlying aging processes in men and women.

## Methods

### Study Design and Participants

The Canadian Longitudinal Study on Aging (CLSA) is a representative, prospective observational study of middle-aged and older adults in Canada that enrolled 51,338 individuals at baseline who were 45 to 85 years of age in 2010 to 2015. The CLSA was designed to be representative of the Canadian population with respect to key demographic characteristics. Participants were recruited through a random, stratified sampling strategy and assigned to one of two groups: the tracking cohort or the comprehensive cohort. A subset of participants from the comprehensive cohort, including 30,097 participants recruited from seven of Canada’s ten provinces, serves as the source population for the present analyses. These individuals lived within 25-50 kilometers of 11 data collection sites across the seven provinces, and available information was obtained through in-person home interviews, tests, physical measurements, and biospecimens (blood and urine).^23^ For the current study, we used baseline data from up to 8,516 CLSA participants to assess the role of adverse socioeconomic conditions on markers of systemic inflammation and biological aging (**Figure 1**).

**Figure 1:**
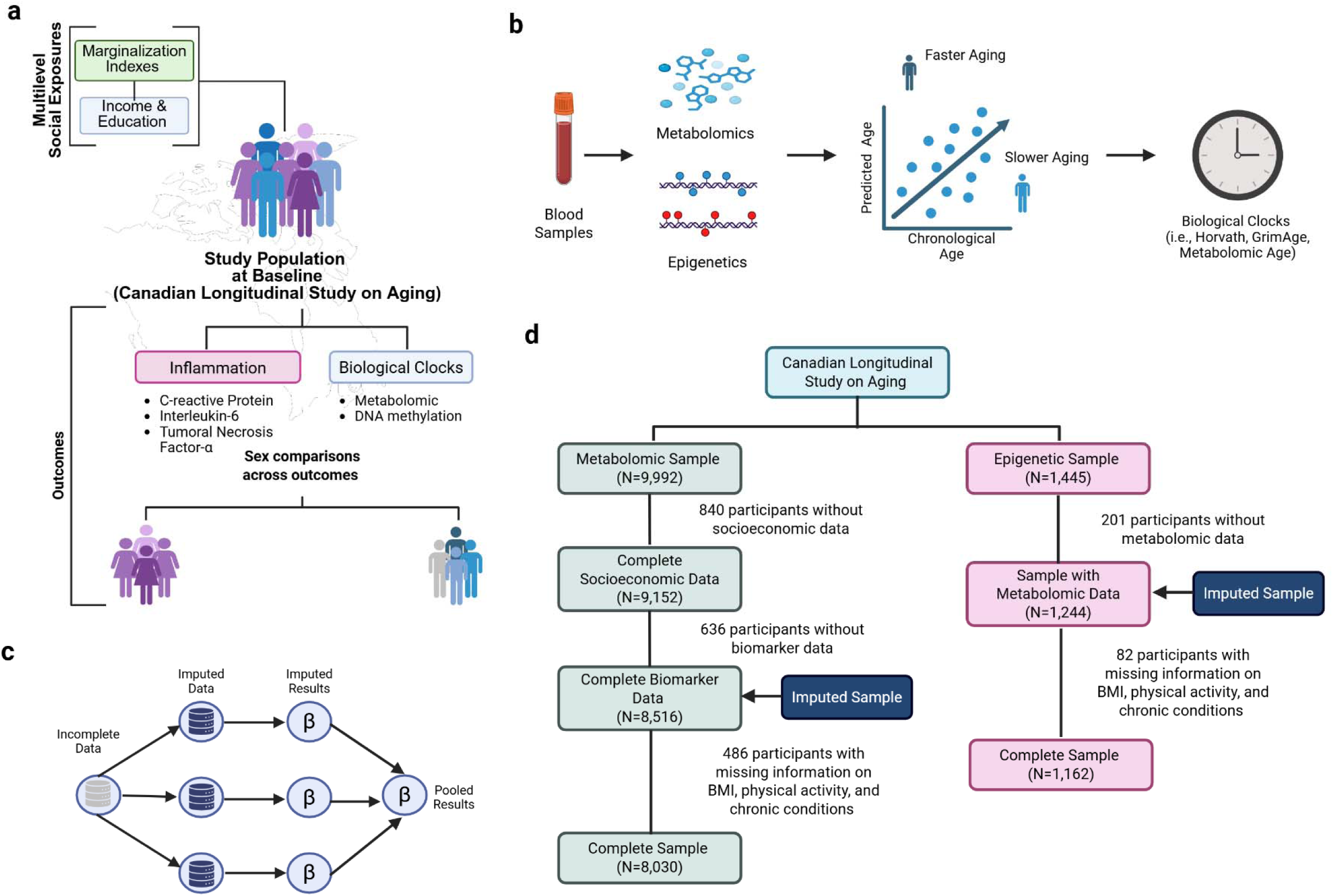
Estimating the association between socioeconomic adversity, systemic inflammation, and biological aging in the Canadian Longitudinal Study on Aging (CLSA). **a)** Representation of cross-sectional analysis of multilevel social exposures and blood-based biomarkers by sex. **b)** For blood-based biomarkers, we estimated metabolomic and epigenetic clocks using linear regression models. **c)** Missing data was imputed following multiple imputation by chained equation (MICE), and pooled estimates were reported. **d)** Flowchart of sample selection for multiple imputation and analysis.

### Measures

#### Outcomes: Inflammatory Markers and Biological Aging

##### Immune Markers of Systemic Inflammation

C-reactive protein (CRP), Interleukin-6 (IL-6), and Tumoral Necrosis Factor-α (TNF-α) were measured from baseline samples and were available for 8,516 participants. Biospecimen collection took place at each data collection site, blood samples were processed within 2 hours after collection, and stored at -80°C before shipping to the CLSA biorepository for storage. Approximately 60mL of non-fasting blood were collected into six tube types to produce serum, plasma, buffy coat two types of peripheral mononuclear cells, and three types of whole blood including dried blood spots.^23^

##### Metabolomic clock and age acceleration

Plasma samples from 9,992 individuals from the CLSA baseline collection underwent metabolomic profiling (Metabolon, Inc, Durham NC), which quantified levels of 1,458 metabolites using Ultrahigh Performance Liquid Chromatography-Tandem Mass Spectroscopy (UPLC-MS/MS). Quality control (QC) and data curation were applied to ensure consistent identification of true chemical entities and to remove those representing systemic artifacts, mis-assignments, and background noise. Data quality assessment was performed using Metabolon internal QC and standards, longitudinal QC samples, batch-based measurements, and methods comparison for selected metabolites. Peaks were quantified using area-under-the-curve, and data normalization was performed to correct for inter-day tuning differences. We used batch-normalized metabolite levels, including metabolites that were bridged across every batch. A detailed description of quality control and normalization process can be found elsewhere.^24^ We then applied a metabolomic age prediction model also developed on Metabolon’s untargeted metabolomics platform from 11,977 persons from the INTERVAL study (age range 18-75 years) and validated in the NEO study (n=599; age range 45-65 years). This prediction clock was developed by applying ridge regression and bootstrapping of 826 metabolites (678 endogenous and 148 xenobiotics).^25^ We used log-transformed metabolite levels to translate metabolomic age estimates to the CLSA, including 811 overlapping metabolites (98%). Metabolite levels may vary by biological sex; as a result, we used the sex specific models for calculating metabolomic age separately for men and women .^25^ We then calculated metabolomic age accelerated residuals using linear models and regressing metabolomic age (Y) on a person’s chronological age (X) given their sex. Residual values greater than zero indicated accelerated metabolomic age, whereas values of less than zero indicated decelerated metabolomic age.

##### DNAm clocks and age acceleration

DNAm data were available for a subsample of 1,445 CLSA participants from baseline samples. Genome-wide DNAm profiling was performed on peripheral blood mononuclear cells (PBMCs), using the Illumina Infinium MethylationEPIC v1 microarray (Illumina, CA, USA). The EPIC array quantitatively measures DNA methylation at 862,927 CpG sites and 2,932 CHH sites across the genome. Data pre-processing included sample- and array-level quality control assessments, probe filtering, outlier detection, normalization, background and batch correction, estimation and adjustment for cell-type composition. A detailed description of the DNA methylation quality control and normalization procedures is available elsewhere.^26^ We then calculated estimates of epigenetic aging from DNAm data using coefficients from six well-calibrated epigenetic clocks: two first-generation (i.e., Horvath, Hannum) clocks, which are highly predictive of chronological age; and four second-generation clocks (PhenoAge, GrimAge, GrimAge2, and Dunedin pace of aging), which are more predictive of physiological states associated with morbidity and mortality. We prioritized these second generation clocks, as they incorporate additional physiological markers of aging and tend to be better predictors of morbidity than first generation clocks.^27^ Similar to metabolomic clocks, we calculated Epigenetic Age Accelerations (EAAs) as a residual of regressing DNAm age (Y) on chronological age (X) while adjusting for DNAm-based estimation of cell type composition (e.g., CD4T, Natural Killer, Monocytes, Plasma Blastocytes, CD8pCD28nCD45Ran, CD8 naïve), which may be a confounder of epigenetic age predictors.^28^ We selected these cell types based on the original selection of the developers of the Horvath clock, and dropped granulocytes from the adjustment set due to the high correlation with other cell types.^29,30^

EAA measures were used in the analysis as a measure of accelerated biological aging. Pace of aging was measured using the DunedinPACE algorithm, which quantifies the current rate of biological aging, with a value of 1.0 corresponding to one year of biological aging per chronological year.

### Exposure

We used 4 different measures of socioeconomic position to assess the role of adversity on biological aging and inflammation, two individual-level measures and two neighborhood level measures. Income was measured at the individual level and used as a categorical variable to denote the following levels <$20K, ≥$20K & <$50K, ≥$50K & <$100K, and ≥$100K (reference). Educational attainment was also measured at the individual level and used as a categorical variable denoting three levels: less than secondary school graduation, secondary school graduation/some post-secondary school graduation, and post-secondary degree or diploma (reference). The material deprivation index was measured at the neighborhood-level and represents a composite measure of three neighborhood characteristics: percentage of people without a high school diploma, average household income, and unemployment rate.^31^ The social deprivation index was measured at the neighborhood-level and encompasses three characteristics: proportion of people separated, divorced, or widowed; proportion of people living alone; and proportion of single parent family.^31,32^ For each of these indexes, positive values indicate more neighborhood material and social deprivation. In analysis, we categorized these indexes in tertiles and quintiles, as well as used them in their continuous form as a sensitivity analysis. Detailed information on how these indexes were calculated can be found elsewhere.^31,32^

### Covariates

We included a set of pre-specified variables as potential confounders that may be associated with socioeconomic adversity, as well as markers of biological aging and systemic inflammation. Demographic factors included age (continuous in years), clinical and behavioral risk factors included body mass index (BMI, calculated as weight in kilograms divided by height in meters squared, continuous), exercise (measured using the physical activity scale for the elderly, continuous),^33^ smoking status (currently, former, never smoker), alcohol drinking (ever drunk alcohol: yes, no), having ≥1 chronic health condition among 38 different long-term physical or mental health conditions including heart disease, kidney disease, chronic obstructive pulmonary disease, asthma, high blood pressure, diabetes, Alzheimer’s, among others. (yes, no).

### Statistical Analysis

Among the 9,992 participants with complete metabolomic age data, we further restricted analyses to individuals with complete information on all exposures and outcomes of interest; 840 participants lacked data on socioeconomic conditions, and 636 lacked biomarker data (**Figure 1**). Our sample data (n=8,516) had 486 (5.7%) participants with missing information in either BMI, physical activity, or chronic health conditions. Baseline characteristics were compared by sex using Welch’s two-sample t-tests for continuous variables and Pearson’s chi-squared tests for categorical variables. Initial analyses also inspected the distribution graphically of metabolomic age acceleration by sex. We fit multivariable-adjusted linear regression models to estimate the association between each one of the 4 exposure variables (income, education, material deprivation, and social deprivation) and metabolomic age acceleration stratified by sex and adjusting for age, BMI, physical activity, smoking status, alcohol consumption, chronic disease, and immune markers (C-reactive protein, Interleukin-6, Tumoral Necrosis Factor-α). We also explored the association between each one of the adverse socioeconomic conditions and log-transformed CRP, IL-6, and TNF-α. These models were stratified by sex and adjusted by the same set of covariates described above, including metabolomic age acceleration. We used multivariate imputation by chained equation (MICE) analysis to account for missingness and obtained pool estimates of our regression models (**Figure 1**).^34,35^ To perform MICE models, we used age, sex, and physical activity to impute BMI; age, sex, and BMI to impute physical activity; and age, sex, BMI, and physical activity to impute chronic health conditions. The variables BMI and physical activity were imputed using predicted mean matching, and chronic health conditions using logistic regression. For each MICE model, we constructed 5 imputed datasets and set a random seed for reproducibility purposes and visually inspected the mean distribution of BMI and physical activity scores. These analyses were conducted using the mice R package.^36^ We also explored the relationship between socioeconomic conditions and epigenetic age acceleration among the subset of individuals with DNA methylation data (n=1,244) across the six pre-specified DNAm clocks. Similarly, we used MICE to obtain pool estimates for our regression models, and adjusted for the same set of covariates as well as immune markers (CRP, IL-6, TNF-α) and cell type composition (CD8 naïve, CD8pCD28nCD45Ran, Plasma Blastocytes, CD4T, Natural Killer, Monocytes).

## Results

### Descriptive analyses

For primary analyses, we included 8,516 baseline participants from the CLSA. On average, participants were 62.6 (10.1) years of age, approximately 50% (n=4,258) were females, and 50% were males (n=4,258); nearly 5.5% had less than secondary school graduation, and 15% earned <$20K annually. Participants had on average younger metabolomic age (56.3; SD = 7.8) (**Table 1**). Men were biologically older than women (**Supplemental Figure 1**), had larger BMI, performed more physical activity, and were more likely to be current smokers. We also noted differences in immune markers by sex. That is, men had lower levels of CRP, comparable levels of IL-6, but higher levels of TNF-α than women (**Table 1**). Lastly, men and women lived in comparable materially deprived neighborhoods, but women lived on average on more socially deprived neighborhoods than men.

**Table 1:** Distribution of baseline covariates by sex in the Canadian Longitudinal Study on Aging (CLSA)

| Characteristic | Overall<br>N = 8,516<sup>1</sup> | Women<br>N = 4,258<sup>1</sup> | Men<br>N = 4,258<sup>1</sup> | p-value<sup>2</sup> |
| --- | --- | --- | --- | --- |
| **Age (years)** | 62.63 (10.09) | 62.29 (10.02) | 62.98 (10.15) | **0.002** |
| **Income** |  |  |  | **<0.001** |
| 1. <\$20K | 1,258 (15%) | 919 (22%) | 339 (8.0%) | |
| 2. ≥20K & <50K | 3,154 (37%) | 1,815 (43%) | 1,339 (31%) |  |
| 3. ≥50K & <100K | 2,925 (34%) | 1,208 (28%) | 1,717 (40%) |  |
| 4. ≥100K | 1,179 (14%) | 316 (7.4%) | 863 (20%) |  |
| **Education** |  |  |  | **<0.001** |
| 1. Less than secondary school graduation | 469 (5.5%) | 264 (6.2%) | 205 (4.8%) |  |
| 2. Secondary school graduation/Some post-secondary education | 1,413 (17%) | 772 (18%) | 641 (15%) |  |
| 3. Post-secondary degree/diploma | 6,634 (78%) | 3,222 (76%) | 3,412 (80%) |  |
| **Residuals** | 0.00 (5.93) | -1.40 (5.23) | 1.40 (6.24) | **<0.001** |
| **Metabolomic Age (years)** | 56.26 (7.82) | 54.68 (6.92) | 57.83 (8.34) | **<0.001** |
| **Body Mass Index (kg/m<sup>2</sup>)** | 28.07 (5.40) | 27.87 (5.94) | 28.27 (4.79) | **<0.001** |
| Missing BMI | 15 | 7 | 8 |  |
| **Physical Activity Score (PSA)** | 143.51 (74.14) | 134.35 (70.34) | 152.63 (76.68) | **<0.001** |
| Missing PSA | 432 | 222 | 210 |  |
| **Smoking Status** |  |  |  | **<0.001** |
| Former | 3,684 (43%) | 1,726 (41%) | 1,958 (46%) |  |
| No, (don't)/never have | 4,029 (47%) | 2,164 (51%) | 1,865 (44%) |  |
| Yes, Currently | 803 (9.4%) | 368 (8.6%) | 435 (10%) |  |
| **Drinking Status** |  |  |  | **<0.001** |
| No | 184 (2.2%) | 117 (2.7%) | 67 (1.6%) |  |
| Yes | 8,332 (98%) | 4,141 (97%) | 4,191 (98%) |  |
| **Chronic Disease** |  |  |  | **<0.001** |
| At least 1 | 7,948 (94%) | 4,020 (95%) | 3,928 (93%) |  |
| No | 522 (6.2%) | 219 (5.2%) | 303 (7.2%) |  |
| Missing Chronic Disease | 46 | 19 | 27 |  |
| **C-reactive Protein (log-scale)** | 0.32 (0.96) | 0.41 (1.00) | 0.22 (0.92) | **<0.001** |
| **Interleukin-6 (log-scale)** | 0.67 (0.63) | 0.67 (0.64) | 0.66 (0.62) | 0.486 |
| **Tumoral Necrosis Factor (log-scale)** | 0.02 (0.35) | -0.01 (0.36) | 0.05 (0.34) | **<0.001** |
| **Material Deprivation Index** | -0.02 (0.04) | -0.02 (0.04) | -0.02 (0.04) | **0.007** |
| **Social Deprivation Index** | 0.00 (0.04) | 0.01 (0.04) | 0.00 (0.04) | **<0.001** |
| **Material Deprivation Index** |  |  |  | 0.171 |
| Quintile 1 | 1,704 (20%) | 836 (20%) | 868 (20%) |  |
| Quintile 2 | 1,703 (20%) | 834 (20%) | 869 (20%) |  |
| Quintile 3 | 1,703 (20%) | 836 (20%) | 867 (20%) |  |
| Quintile 4 | 1,703 (20%) | 857 (20%) | 846 (20%) |  |
| Quintile 5 | 1,703 (20%) | 895 (21%) | 808 (19%) |  |
| **Social Deprivation Index** |  |  |  | **<0.001** |
| Quintile 1 | 1,704 (20%) | 785 (18%) | 919 (22%) |  |
| Quintile 2 | 1,703 (20%) | 824 (19%) | 879 (21%) |  |
| Quintile 3 | 1,703 (20%) | 845 (20%) | 858 (20%) |  |
| Quintile 4 | 1,703 (20%) | 893 (21%) | 810 (19%) |  |
| Quintile 5 | 1,703 (20%) | 911 (21%) | 792 (19%) |  |

<sup>1</sup>Mean (SD); n (%)<sup>2</sup>Welch Two Sample t-test; Pearson's Chi-squared test

We next examined correlations between inflammatory biomarkers and biological clocks in men and women within a subsample of the CLSA (n = 1,244). Epigenetic clocks were moderately and similarly correlated in women (median = 0.62, IQR = 0.12) and men (median = 0.61, IQR = 0.13). Correlations among inflammatory biomarkers were also similar between women (median = 0.40, IQR = 0.15) and men (median = 0.36, IQR = 0.12). EAA residuals from second-generation clocks showed moderate correlations with CRP and IL-6 in both men and women. They also exhibited low correlation with metabolomic age, indicating that epigenetic and metabolomic clocks likely measure different and complementary biological processes. Lastly, metabolomic age acceleration measures were also correlated with IL-6 and TNF-α, although these associations were modest (**Figure 2**).

**Figure 2:**
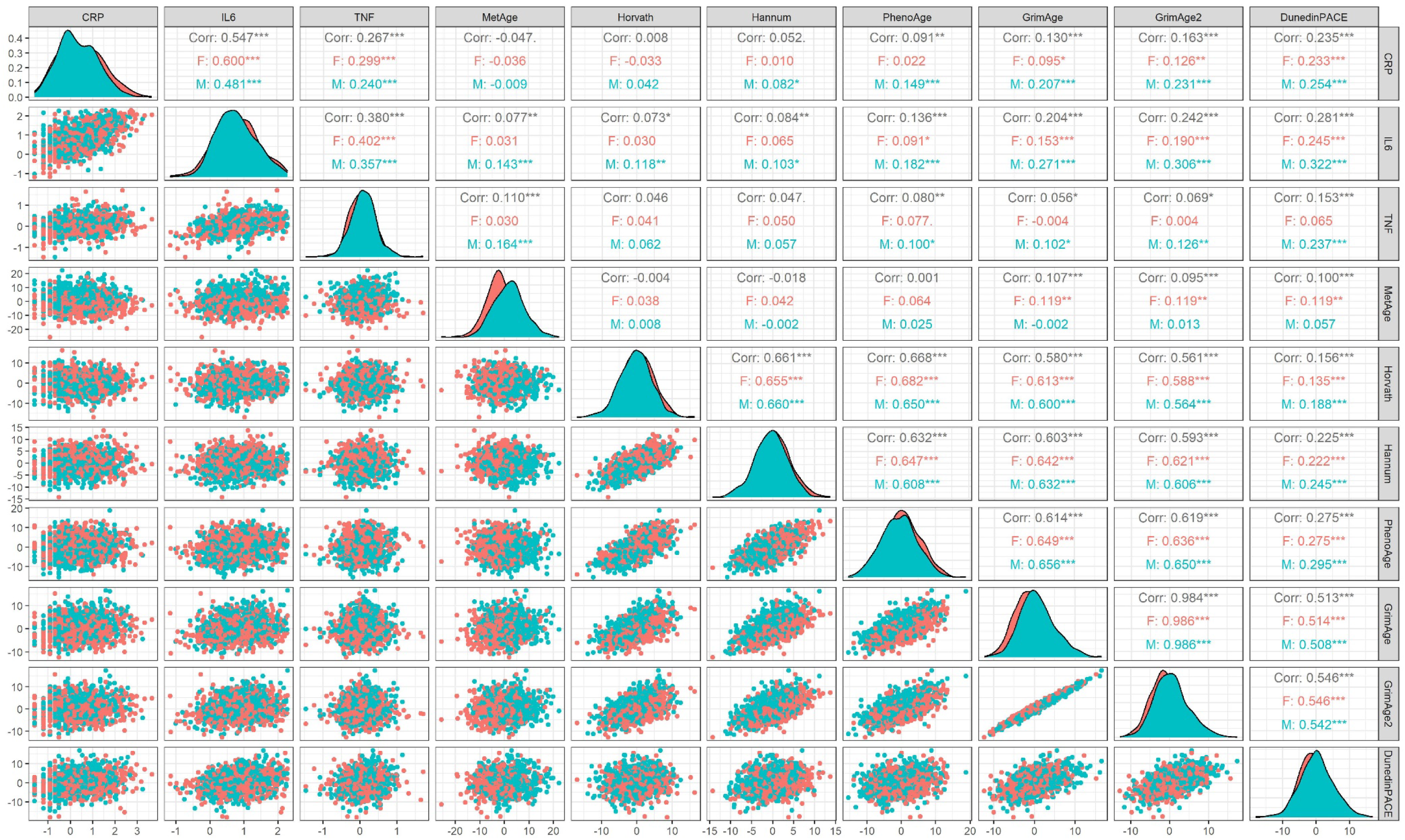
Correlation plot between systemic inflammation markers, metabolomic age clock, and DNA methylation clocks by males and females in the Canadian Longitudinal Study on Aging (CLSA, n=1,244)

### Association between socioeconomic conditions and metabolomic age acceleration

In multivariable linear adjusted models, we found that in comparison to those earning ≥$100K a year, women earning less <$20K were on average 1.14 (95%CI: 0.46, 1.82) year older with respect to metabolomic age; those earning ≥$20K & <$50K were on average 0.90 (95%CI: 0.26, 1.53) years older; and those earning ≥$50K & <$100K were on average 0.70 (95%CI: 0.05, 1.34) years older (**Table 2**). We did not observe this dose response among men. Education and neighborhood material deprivation were not associated with metabolomic age acceleration in women. In contrast, among men, greater material deprivation was associated with lower age acceleration (β=-5.30, 95%CI: -10.38, -0.22) (**Table 2**). Among women, residence in highly socially deprived neighborhoods (upper quintile) was associated with 0.55 years greater age acceleration (95% CI: 0.05, 1.05), and each unit increase in social deprivation was associated with 4.97 years greater age acceleration (95% CI: 1.14, 8.8).

**Table 2:**
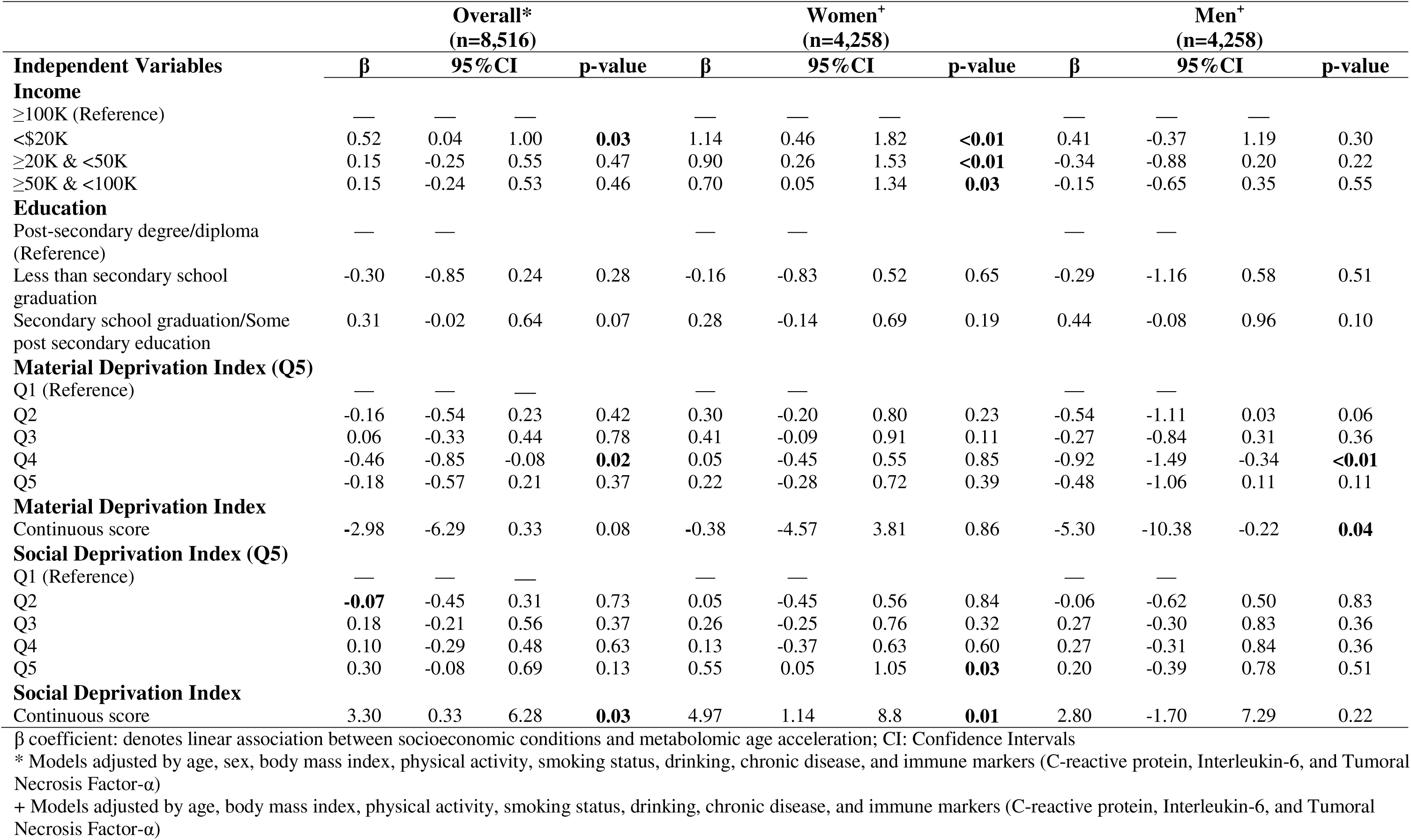
Association between metabolomic age acceleration and socioeconomic conditions stratified by sex in the Canadian Longitudinal Study on Aging (CLSA)

| Independent Variables | Overall*<br>(n=8,516) |  |  |  | Women<sup>+</sup><br>(n=4,258) |  |  |  | Men<sup>+</sup><br>(n=4,258) |  |  |  |
| --- | --- | --- | --- | --- | --- | --- | --- | --- | --- | --- | --- | --- |
| | $\beta$ | 95%CI | p-value | | $\beta$ | 95%CI | p-value | | $\beta$ | 95%CI | p-value | |
| **Income** |  |  |  |  |  |  |  |  |  |  |  |  |
| ≥100K (Reference) | — | — | — |  | — | — | — |  | — | — | — |  |
| <\$20K | 0.52 | 0.04 | 1.00 | **0.03** | 1.14 | 0.46 | 1.82 | **<0.01** | 0.41 | -0.37 | 1.19 | 0.30 |
| ≥20K & <50K | 0.15 | -0.25 | 0.55 | 0.47 | 0.90 | 0.26 | 1.53 | **<0.01** | -0.34 | -0.88 | 0.20 | 0.22 |
| ≥50K & <100K | 0.15 | -0.24 | 0.53 | 0.46 | 0.70 | 0.05 | 1.34 | **0.03** | -0.15 | -0.65 | 0.35 | 0.55 |
| **Education** |  |  |  |  |  |  |  |  |  |  |  |  |
| Post-secondary degree/diploma (Reference) | — | — |  |  | — | — |  |  | — | — |  |  |
| Less than secondary school graduation | -0.30 | -0.85 | 0.24 | 0.28 | -0.16 | -0.83 | 0.52 | 0.65 | -0.29 | -1.16 | 0.58 | 0.51 |
| Secondary school graduation/Some post secondary education | 0.31 | -0.02 | 0.64 | 0.07 | 0.28 | -0.14 | 0.69 | 0.19 | 0.44 | -0.08 | 0.96 | 0.10 |
| **Material Deprivation Index (Q5)** |  |  |  |  |  |  |  |  |  |  |  |  |
| Q1 (Reference) | — | — | — |  | — | — |  |  | — | — |  |  |
| Q2 | -0.16 | -0.54 | 0.23 | 0.42 | 0.30 | -0.20 | 0.80 | 0.23 | -0.54 | -1.11 | 0.03 | 0.06 |
| Q3 | 0.06 | -0.33 | 0.44 | 0.78 | 0.41 | -0.09 | 0.91 | 0.11 | -0.27 | -0.84 | 0.31 | 0.36 |
| Q4 | -0.46 | -0.85 | -0.08 | **0.02** | 0.05 | -0.45 | 0.55 | 0.85 | -0.92 | -1.49 | -0.34 | **<0.01** |
| Q5 | -0.18 | -0.57 | 0.21 | 0.37 | 0.22 | -0.28 | 0.72 | 0.39 | -0.48 | -1.06 | 0.11 | 0.11 |
| **Material Deprivation Index** |  |  |  |  |  |  |  |  |  |  |  |  |
| Continuous score | -2.98 | -6.29 | 0.33 | 0.08 | -0.38 | -4.57 | 3.81 | 0.86 | -5.30 | -10.38 | -0.22 | **0.04** |
| **Social Deprivation Index (Q5)** |  |  |  |  |  |  |  |  |  |  |  |  |
| Q1 (Reference) | — | — | — |  | — | — |  |  | — | — |  |  |
| Q2 | **-0.07** | -0.45 | 0.31 | 0.73 | 0.05 | -0.45 | 0.56 | 0.84 | -0.06 | -0.62 | 0.50 | 0.83 |
| Q3 | 0.18 | -0.21 | 0.56 | 0.37 | 0.26 | -0.25 | 0.76 | 0.32 | 0.27 | -0.30 | 0.83 | 0.36 |
| Q4 | 0.10 | -0.29 | 0.48 | 0.63 | 0.13 | -0.37 | 0.63 | 0.60 | 0.27 | -0.31 | 0.84 | 0.36 |
| Q5 | 0.30 | -0.08 | 0.69 | 0.13 | 0.55 | 0.05 | 1.05 | **0.03** | 0.20 | -0.39 | 0.78 | 0.51 |
| **Social Deprivation Index** |  |  |  |  |  |  |  |  |  |  |  |  |
| Continuous score | 3.30 | 0.33 | 6.28 | **0.03** | 4.97 | 1.14 | 8.8 | **0.01** | 2.80 | -1.70 | 7.29 | 0.22 |

$\beta$ coefficient: denotes linear association between socioeconomic conditions and metabolomic age acceleration; CI: Confidence Intervals
\* Models adjusted by age, sex, body mass index, physical activity, smoking status, drinking, chronic disease, and immune markers (C-reactive protein, Interleukin-6, and Tumoral Necrosis Factor-α )
+ Models adjusted by age, body mass index, physical activity, smoking status, drinking, chronic disease, and immune markers (C-reactive protein, Interleukin-6, and Tumoral Necrosis Factor-α )

### Association between socioeconomic conditions and immune markers of systemic inflammation

In multivariable-adjusted models, lower income was associated with higher circulating IL-6 levels in both women and men. Compared with those earning ≥$100K annually, women earning <$20K, ≥$20-50K, and ≥$50-100K had 10% (95% CI: 2%, 17%), 6% (95% CI: 0%, 14%), and 8% (95% CI: 1%, 15%) higher IL-6 levels, respectively (**Table 3**). Corresponding increases among men were 20% (95% CI: 12%, 28%), 15% (95% CI: 10%, 21%), and 5% (95% CI: 1%, 10%; **Table 4**). Although men had lower circulating CRP levels than women, lower income was associated with higher CRP levels among men but not women (**Tables 3 and 4**).

**Table 3:**
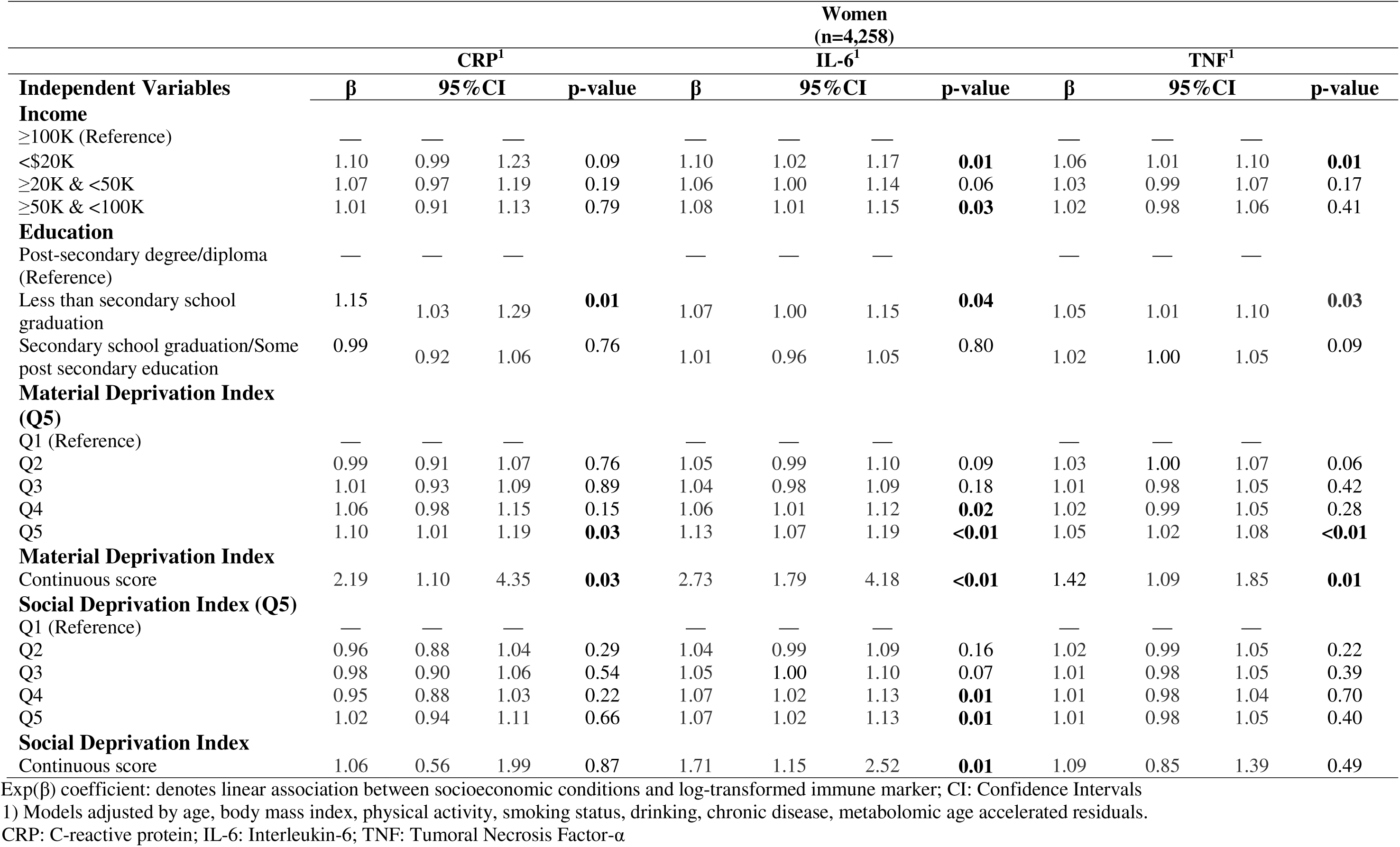
Association between immune markers of systemic inflammation and socioeconomic conditions among women of the Canadian Longitudinal Study on Aging (CLSA).

**Table 4:** Association between immune markers of systemic inflammation and socioeconomic conditions among men of the Canadian Longitudinal Study on Aging (CLSA).

| Independent Variables | Men<br>(n=4,258) |  |  |  |  |  |  |  |  |  |  |  |
| --- | --- | --- | --- | --- | --- | --- | --- | --- | --- | --- | --- | --- |
|  | CRP<sup>1</sup> |  |  |  | IL-6<sup>1</sup> |  |  |  | TNF<sup>1</sup> |  |  |  |
| | $\beta$ | 95%CI | p-value | | $\beta$ | 95%CI | p-value | | $\beta$ | 95%CI | p-value | |
| **Income** |  |  |  |  |  |  |  |  |  |  |  |  |
| ≥100K (Reference) | — | — | — |  | — | — | — |  | — | — | — |  |
| <\$20K | 1.17 | 1.05 | 1.30 | <0.01 | 1.20 | 1.12 | 1.28 | <0.01 | 1.04 | 1.00 | 1.09 | 0.04 |
| ≥20K & <50K | 1.12 | 1.04 | 1.21 | <0.01 | 1.15 | 1.10 | 1.21 | <0.01 | 1.04 | 1.01 | 1.06 | 0.02 |
| ≥50K & <100K | 1.05 | 0.98 | 1.12 | 0.22 | 1.05 | 1.01 | 1.10 | 0.03 | 1.02 | 0.99 | 1.04 | 0.199 |
| **Education** |  |  |  |  |  |  |  |  |  |  |  |  |
| Post-secondary degree/diploma (Reference) | — | — | — |  | — | — | — |  | — | — | — |  |
| Less than secondary school graduation | 1.15 | 1.02 | 1.29 | 0.03 | 1.12 | 1.04 | 1.21 | <0.01 | 1.09 | 1.04 | 1.14 | <0.01 |
| Secondary school graduation/Some post secondary education | 1.02 | 0.95 | 1.10 | 0.54 | 1.05 | 1.00 | 1.10 | 0.05 | 1.01 | 0.98 | 1.04 | 0.43 |
| **Material Deprivation Index (Q5)** |  |  |  |  |  |  |  |  |  |  |  |  |
| Q1 (Reference) | — | — | — |  | — | — | — |  | — | — | — |  |
| Q2 | 1.05 | 0.97 | 1.14 | 0.24 | 1.04 | 0.99 | 1.09 | 0.16 | 1.03 | 1.00 | 1.06 | 0.08 |
| Q3 | 1.02 | 0.94 | 1.10 | 0.72 | 1.05 | 1.00 | 1.10 | 0.05 | 1.01 | 0.98 | 1.04 | 0.56 |
| Q4 | 1.11 | 1.03 | 1.21 | <0.01 | 1.11 | 1.06 | 1.17 | <0.01 | 1.04 | 1.01 | 1.07 | 0.01 |
| Q5 | 1.17 | 1.08 | 1.27 | <0.01 | 1.16 | 1.10 | 1.22 | <0.01 | 1.05 | 1.02 | 1.08 | <0.01 |
| **Material Deprivation Index Continuous score** |  |  |  |  |  |  |  |  |  |  |  |  |
| **Social Deprivation Index (Q5)** | 4.50 | 2.23 | 9.06 | <0.01 | 4.18 | 2.69 | 6.50 | <0.01 | 1.53 | 1.17 | 1.99 | <0.01 |
| Q1 (Reference) | — | — | — |  | — | — | — |  | — | — | — |  |
| Q2 | 1.10 | 1.02 | 1.19 | 0.01 | 1.05 | 1.00 | 1.10 | 0.06 | 1.02 | 0.99 | 1.05 | 0.20 |
| Q3 | 1.12 | 1.04 | 1.21 | <0.01 | 1.05 | 1.00 | 1.10 | 0.06 | 0.99 | 0.97 | 1.02 | 0.67 |
| Q4 | 1.15 | 1.06 | 1.24 | <0.01 | 1.11 | 1.06 | 1.17 | <0.01 | 1.02 | 0.99 | 1.05 | 0.16 |
| Q5 | 1.12 | 1.04 | 1.22 | 0.01 | 1.11 | 1.05 | 1.16 | <0.01 | 1.04 | 1.01 | 1.07 | 0.01 |
| **Social Deprivation Index Continuous score** | 1.89 | 1.02 | 3.53 | 0.04 | 2.35 | 1.59 | 3.49 | <0.01 | 1.35 | 1.07 | 1.70 | 0.01 |

Exp( $\beta$ ) coefficient: denotes linear association between socioeconomic conditions and log-transformed immune marker; CI: Confidence Intervals 1) Models adjusted by age, body mass index, physical activity, smoking status, drinking, chronic disease, metabolomic age accelerated residuals. CRP: C-reactive protein; IL-6: Interleukin-6; TNF: Tumoral Necrosis Factor-α

Among women, residence in the highest quintile of material deprivation was associated with higher CRP, IL-6, and TNF-α levels, while social deprivation was associated only with higher IL-6 (**Table 3**). Among men, those living in the fourth and highest quintile of material and social deprivation had similarly elevated levels of inflammatory markers (**Table 4**). Among women, having less than secondary school education (vs. post-secondary degree or diploma) was associated with higher circulating levels of CRP (β=15%; 95% CI: 3%, 29%), IL-6 (β=7%; 95% CI: 0%, 15%), and TNF-α (β=5%; 95% CI: 1%, 10%) (**Table 3**). In men, lower educational attainment was also associated with these inflammatory biomarkers in a similar way. (**Table 4**).

### Association between socioeconomic conditions and epigenetic age acceleration

We confirmed approximate normality of EAA residuals (**Supplemental Figures 2 and 3**). Among women, residence in the highest tertile of material deprivation (vs. lowest) was associated with greater age acceleration across all second-generation clocks, including GrimAge (β=0.90 years; 95% CI: 0.15, 1.66), GrimAge2 (β=0.98 years; 95% CI: 0.22, 1.75), and Dunedin Pace of Aging (β=1.72 years; 95% CI: 0.87, 2.57) (**Supplemental Table 1**). Similar patterns were observed for social deprivation, with corresponding increases of 1.34 (95% CI: 0.60, 2.08), 1.33 (95% CI: 0.58, 2.07), and 1.07 (95% CI: 0.23, 1.91) years, respectively. Because Dunedin Pace of Aging indicates the rate of aging per year (rather than years of age acceleration), the prior estimate can be interpreted as a 72% increase in the rate of aging for a person living in highest tertile of material deprivation with respect to a person living in the lowest.

In contrast, income and education were not generally associated with epigenetic age acceleration in women. Among men, income was associated with Dunedin Pace of Aging, but no consistent associations were observed between socioeconomic conditions and other second-generation clocks (**Supplemental Table 2**). In secondary analyses of first-generation epigenetic clocks, social deprivation was associated with Horvath, Hannum, and PhenoAge age acceleration among women (**Supplemental Table 3**). In men, the only corresponding association was between social deprivation and Horvath age acceleration (**Supplemental Table 4**).

## Discussion

Biological aging and systemic inflammation are candidate physiological mechanisms by which socioeconomic adversity may affect human health.^6,15,37–40^ Here, in this cross-sectional analysis of a large, population-based cohort of older adults, socioeconomic factors demonstrated sex-specific associations with biological aging: income was associated with metabolomic age acceleration, and material deprivation with epigenetic age acceleration (e.g., GrimAge, GrimAge2, and Dunedin Pace of Aging), among women but not men. Exposure to lower income and greater material and social deprivation was associated with elevated IL-6 levels in both women and men. However, among men, lower income and higher deprivation were also associated with elevated CRP levels, and TNF-α was associated with higher levels of material deprivation. In contrast, these patterns were less consistent in women, with associations observed primarily between material deprivation and CRP and TNF-α. These findings suggest that socioeconomic adversity influences not only multiple inflammatory pathways but also multi-factorial biological aging processes, including metabolomic and epigenetic aging. The broad impact of these exposures across physiological systems supports their role as potential mechanisms linking the social environment to health and disease. The observed sex differences may reflect both differential exposure to socioeconomic conditions and interactions with underlying biological differences in sex-specific physiology.

A growing body of evidence has demonstrated that socioeconomic adversity influences inflammation and biological aging, and these molecular signatures may impact disease trajectories.^5,6,41^ A recent study of 18 different cohorts spanning 12 countries and exploring the association between low education, as a proxy for socioeconomic position, and first generation epigenetic aging clocks (Horvath, Hannum, and Levine) showed that low education was an independent predictor of epigenetic age acceleration.^42^ For example, relative to those with higher levels of education, lower education levels was associated with 0.20 to 0.60 years epigenetic age acceleration, depending on the specific clock.^42^ We generally did not observe consistent associations between educational attainment and first-generation epigenetic clocks (e.g., Horvath, Hannum). In our study, socioeconomic adversity generally showed stronger and more consistent associations with second-generation epigenetic clocks (GrimAge, GrimAge2, Dunedin) and, in the case of income, with metabolomic age acceleration. These findings suggest that social adversity may have stronger effects on biological aging measures that may reflect health-related phenotypes and may better capture the influence of social determinants on disease trajectories. In a study of approximately 5,000 individuals from three independent cohorts in Italy, Australia, and Ireland, education, used as a marker of socioeconomic position, was associated with epigenetic age acceleration. In pooled analyses, individuals of lower socioeconomic position exhibited nearly one year greater Hannum age acceleration (β=0.99 years; 95% CI: 0.39, 1.59), suggesting that cumulative socioeconomic adversity may become biologically embedded through pathways that accelerate cellular aging.^43^ Another study based in the US, including the Multi-Ethnic Study of Atherosclerosis (MESA, n=1,211) and the Health and Retirement Study (HRS, n=4,018) cohorts, analyzed the relationship between an six-indicator index of socioeconomic position (higher values indicated more socioeconomic disadvantage) and eight epigenetic clocks.^44^ Similar to our findings, second generation clocks such as Dudendin Pace of Aging and GrimAge were strongly positively associated with socioeconomic disadvantage.^44^ These findings, combined with our own, reiterate that biological changes may result from persistent exposure to socioeconomic adversity. Specifically, they reinforce the idea that epigenetic measures may capture downstream effects of cumulative social disadvantage.^43,43–46^

More recently, multi-omics approaches, including metabolomics, have expanded the ability to characterize biological aging. In our study, income was associated with progressively greater metabolomic age acceleration among women, independent of behavioral factors, comorbidities, and immune markers. This result aligns with evidence from a study of individuals with multiple sclerosis and healthy people, where greater area-level deprivation was associated with increased metabolomic age acceleration.^47^ Collectively, these findings suggest that metabolomic measures may complement epigenetic clocks in capturing the biological consequences of socioeconomic conditions.

Notably, we observed sex differences. Despite having lower overall metabolomic age, women exhibited greater acceleration in response to lower income, whereas associations between deprivation and epigenetic aging were also more pronounced in women. These patterns may reflect the unequal distribution of socioeconomic disadvantage, as women are disproportionately represented in lower income strata. In contrast, men exhibited stronger associations between adverse socioeconomic conditions and inflammatory markers, suggesting that different biological systems may capture the effects of social adversity in men and women. Thus, the importance of considering sex-specific pathways linking socioeconomic conditions to biological aging is likely to be relevant.

Systemic inflammation plays a crucial role in the development of chronic conditions that lead to premature mortality.^3^ This link is grounded in modern socio-epidemiological theories that hypothesize that stressful circumstances lead to hyperactivation of the hypothalamic-pituitary-adrenal (HPA) axis, resulting in the release of cortisol and acute immune markers into the bloodstream. This sustained immune stress response could influence biological aging and increase individuals’ susceptibility to chronic diseases, and premature mortality.^48,49^ There is strong evidence that detrimental socioeconomic conditions are associated with alterations in multiple immune stress biomarkers, contributing to an elevated inflammatory response with direct repercussions for morbidity and mortality.^50^ A meta-analysis conducted in North America, including 43 independent studies, demonstrated an association between low socioeconomic position and high levels of CRP and IL-6.^6^ Similar to our findings, evidence from the Health, Aging and Body Composition study, which includes high-functioning older adults (70-79 years old), also corroborated the link between multiple indicators of socioeconomic condition including income, education, and financial assets and markers of systemic inflammation (CRP, IL-6, and TNF-α).^4^ These associations were consistent after excluding participants with chronic conditions, suggesting that the effect of socioeconomic adversity on inflammation is not influenced by prior diseases. Furthermore, longitudinal studies also suggest that neighborhood contextual factors, such as high deprivation were associated with higher CRP levels over time.^51^ Collectively, these findings suggest that socioeconomic adversity may contribute to a pro-inflammatory state characterized by elevated levels of peripheral cytokines that have downstream effects on multiple cellular processes.

Although a growing body of evidence demonstrates consistent associations among socioeconomic adversity, biological aging, and systemic inflammation, these studies have generally overlooked sex-dependent patterns in their links to inflammatory response or aging. Our sex-stratified analyses highlight the importance of examining these associations separately, as income-related disadvantage showed distinct relationships with metabolomic aging among women and inflammatory biomarkers among men. Exploring these patterns requires careful consideration in future research, as they may reflect distinct sex-specific biological pathways through which socioeconomic adversity influences health and disease.

This study has several limitations. First, our sample is cross-sectional, which limits our ability to make causal inferences and precludes us from investigating how social conditions affect trajectories of biological aging and systemic inflammation. Although our independent variables include two individual-level indicators of socioeconomic position (i.e., income and education) and two neighborhood-level indicators of socioeconomic disadvantage (i.e., the social deprivation and material deprivation indices), our analysis does not include childhood measures of socioeconomic adversity. Thus, we are unable to assess how exposure to adverse social conditions in early life affects biological pathways in late adulthood. Lastly, despite adjusting for common pro-inflammatory risk factors (e.g., smoking and alcohol consumption) and chronic health conditions, the observational nature of our study design makes this analysis susceptible to residual confounding.

Our study also has several strengths, including a large sample of older adults with information on important covariates of interest. We characterized the effects of our socioeconomic indicators while controlling for major behavioral risk factors and chronic conditions, thereby attenuating potential confounding bias. Our study incorporated several biological aging clocks to capture distinct aspects of aging, as well as multiple biomarkers of systemic inflammation, allowing us to examine different pathways through which social adversity may become biologically embodied. Future research should explore how these processes are interconnected and jointly influence human health.

In conclusion, we demonstrated that exposure to detrimental socioeconomic conditions at the individual and neighborhood levels are associated with specific measures of biological aging in women, but not in men. Sex-dependent patterns were also observed for systemic inflammation; we found that men are more likely to exhibit higher levels of systemic inflammation when exposed to lower income and high material and social deprivation. Systemic inflammation has been documented as a potential causal pathway for health inequities in chronic conditions.^52^ We suggest that future research continue to explore sex-dependent patterns between social exposures and biological mechanisms of health and disease. Temporal data might help elucidate the connections between these interrelated biological processes.

## Authors’ Approval

All authors have seen and approved the submission of this manuscript.

## Competing Interest

None

## Data Availability

Data are available from the Canadian Longitudinal Study on Aging (www.clsa-elcv.ca) for researchers who meet the criteria for access to de-identified CLSA data.

## Funding Information

Cesar Higgins Tejera was supported by the Alzheimer’s Association Research Fellowship to Promote Diversity (AARF-D) (grant number AARFD-24-1306587).

**Supplemental Table 1:** Association between DNA methylation age acceleration and socioeconomic conditions among female participants of the Canadian Longitudinal Study on Aging (CLSA)

| Independent Variables | Women (n=624) |  |  |  |  |  |  |  |  |  |  |  |
| --- | --- | --- | --- | --- | --- | --- | --- | --- | --- | --- | --- | --- |
|  | Grim<sup>1</sup> |  |  |  | Grim2<sup>1</sup> |  |  |  | Dunedin<sup>1</sup> |  |  |  |
| | $\beta$ | 95%CI | p-value | | $\beta$ | 95%CI | p-value | | $\beta$ | 95%CI | p-value | |
| **Income** |  |  |  |  |  |  |  |  |  |  |  |  |
| ≥100K (Reference) | — | — | — |  | — | — | — |  | — | — | — |  |
| <\$20K | 0.52 | -0.75 | 1.78 | 0.43 | 0.69 | -0.58 | 1.97 | 0.29 | 0.85 | -0.57 | 2.28 | 0.24 |
| ≥20K & <50K | 0.11 | -1.09 | 1.30 | 0.86 | 0.21 | -0.99 | 1.41 | 0.73 | 0.03 | -1.31 | 1.37 | 0.97 |
| ≥50K & <100K | -0.26 | -1.48 | 0.96 | 0.68 | -0.18 | -1.41 | 1.05 | 0.77 | -0.45 | -1.82 | 0.92 | 0.52 |
| **Education** |  |  |  |  |  |  |  |  |  |  |  |  |
| Post-secondary degree/diploma (Reference) | — | — | — |  | — | — | — |  | — | — | — |  |
| Less than secondary school graduation | 0.44 | -0.80 | 1.69 | 0.49 | 0.29 | -0.97 | 1.54 | 0.66 | 1.17 | -0.23 | 2.57 | 0.10 |
| Secondary school graduation/Some post secondary education | -0.11 | -0.91 | 0.68 | 0.78 | 0.03 | -0.77 | 0.83 | 0.94 | -0.12 | -1.02 | 0.77 | 0.79 |
| **Material Deprivation Index (T3)** |  |  |  |  |  |  |  |  |  |  |  |  |
| T1 (Reference) | — | — | — |  | — | — | — |  | — | — | — |  |
| T2 | 0.18 | -0.56 | 0.93 | 0.63 | 0.24 | -0.51 | 0.99 | 0.53 | 0.55 | -0.27 | 1.38 | 0.19 |
| T3 | 0.90 | 0.15 | 1.66 | **0.02** | 0.98 | 0.22 | 1.75 | **0.01** | 1.72 | 0.87 | 2.57 | **<0.01** |
| **Material Deprivation Index** |  |  |  |  |  |  |  |  |  |  |  |  |
| Continuous score | 10.15 | 2.37 | 17.94 | **0.01** | 11.05 | 3.22 | 18.89 | **0.01** | 19.83 | 11.17 | 28.49 | **<0.01** |
| **Social Deprivation Index (T3)** |  |  |  |  |  |  |  |  |  |  |  |  |
| T1 (Reference) | — | — | — |  | — | — | — |  | — | — | — |  |
| T2 | 1.58 | 0.84 | 2.31 | **<0.01** | 1.55 | 0.81 | 2.30 | **<0.01** | 0.79 | -0.05 | 1.63 | 0.06 |
| T3 | 1.34 | 0.60 | 2.08 | **<0.01** | 1.33 | 0.58 | 2.07 | **<0.01** | 1.07 | 0.23 | 1.91 | **0.01** |
| **Social Deprivation Index** |  |  |  |  |  |  |  |  |  |  |  |  |
| Continuous score | 14.61 | 7.02 | 22.21 | **<0.01** | 14.26 | 6.61 | 21.92 | **<0.01** | 10.32 | 1.71 | 18.93 | **0.02** |

$\beta$ coefficient: denotes linear association between socioeconomic conditions and epigenetic age acceleration; CI: Confidence Intervals
1) Models adjusted by age, body mass index, physical activity, smoking status, drinking, chronic disease, immune markers (i.e., C-reactive protein, Interleukin-6, Tumoral Necrosis Factor-α ) and cell types (i.e., CD4T, Natural Killer, Monocytes, Plasma Blastocytes, CD8pCD28nCD45Ran, CD8 naïve)

**Supplemental Table 2:** Association between DNA methylation age acceleration and socioeconomic conditions among male participants of the Canadian Longitudinal Study of Aging (CLSA)

| Independent Variables | Men (n=620) |  |  |  |  |  |  |  |  |  |  |  |
| --- | --- | --- | --- | --- | --- | --- | --- | --- | --- | --- | --- | --- |
|  | Grim<sup>1</sup> |  |  |  | Grim2<sup>1</sup> |  |  |  | Dunedin<sup>1</sup> |  |  |  |
| | $\beta$ | 95%CI | p-value | | $\beta$ | 95%CI | p-value | | $\beta$ | 95%CI | p-value | |
| **Income** |  |  |  |  |  |  |  |  |  |  |  |  |
| ≥100K (Reference) | — | — | — |  | — | — | — |  | — | — | — |  |
| <\$20K | 0.77 | -0.39 | 1.93 | 0.19 | 0.90 | -0.26 | 2.06 | 0.13 | 1.78 | 0.41 | 3.16 | **0.01** |
| ≥20K & <50K | 0.29 | -0.63 | 1.22 | 0.54 | 0.35 | -0.57 | 1.27 | 0.46 | 0.55 | -0.54 | 1.65 | 0.32 |
| ≥50K & <100K | -0.10 | -0.92 | 0.71 | 0.80 | -0.14 | -0.96 | 0.67 | 0.73 | 0.04 | -0.92 | 1.01 | 0.93 |
| **Education** |  |  |  |  |  |  |  |  |  |  |  |  |
| Post-secondary degree/diploma (Reference) | — | — | — |  | — | — | — |  | — | — | — |  |
| Less than secondary school graduation | 1.88 | 0.44 | 3.32 | **0.01** | 1.96 | 0.52 | 3.40 | **0.01** | 1.33 | -0.39 | 3.04 | 0.13 |
| Secondary school graduation/Some post secondary education | -0.20 | -1.04 | 0.64 | 0.64 | -0.30 | -1.13 | 0.54 | 0.49 | 0.69 | -0.31 | 1.69 | 0.18 |
| **Material Deprivation Index (T3)** |  |  |  |  |  |  |  |  |  |  |  |  |
| T1 (Reference) | — | — | — |  | — | — | — |  | — | — | — |  |
| T2 | 0.13 | -0.61 | 0.86 | 0.74 | 0.19 | -0.54 | 0.93 | 0.61 | -0.42 | -1.29 | 0.46 | 0.35 |
| T3 | -0.15 | -0.91 | 0.61 | 0.69 | -0.13 | -0.89 | 0.63 | 0.73 | -0.07 | -0.97 | 0.83 | 0.87 |
| **Material Deprivation Index** |  |  |  |  |  |  |  |  |  |  |  |  |
| Continuous score | 0.03 | -7.71 | 7.76 | 0.99 | -0.17 | -7.9 | 7.56 | 0.97 | -1.43 | -10.61 | 7.76 | 0.76 |
| **Social Deprivation Index (T3)** |  |  |  |  |  |  |  |  |  |  |  |  |
| T1 (Reference) | — | — | — |  | — | — | — |  | — | — | — |  |
| T2 | 0.31 | -0.43 | 1.05 | 0.41 | 0.34 | -0.40 | 1.08 | 0.37 | -0.19 | -1.07 | 0.69 | 0.68 |
| T3 | 0.55 | -0.20 | 1.30 | 0.15 | 0.42 | -0.32 | 1.17 | 0.27 | -0.28 | -1.17 | 0.61 | 0.54 |
| **Social Deprivation Index** |  |  |  |  |  |  |  |  |  |  |  |  |
| Continuous score | 6.43 | -0.87 | 13.73 | 0.08 | 5.76 | -1.55 | 13.06 | 0.12 | -1.49 | -10.18 | 7.2 | 0.74 |

$\beta$ coefficient: denotes linear association between socioeconomic conditions and epigenetic age acceleration; CI: Confidence Intervals
1) Models adjusted by age, body mass index, physical activity, smoking status, drinking, chronic disease, immune markers (i.e., C-reactive protein, Interleukin-6, Tumoral Necrosis Factor-α ) and cell types (i.e., CD4T, Natural Killer, Monocytes, Plasma Blastocytes, CD8pCD28nCD45Ran, CD8 naïve)

**Supplemental Table 3:**
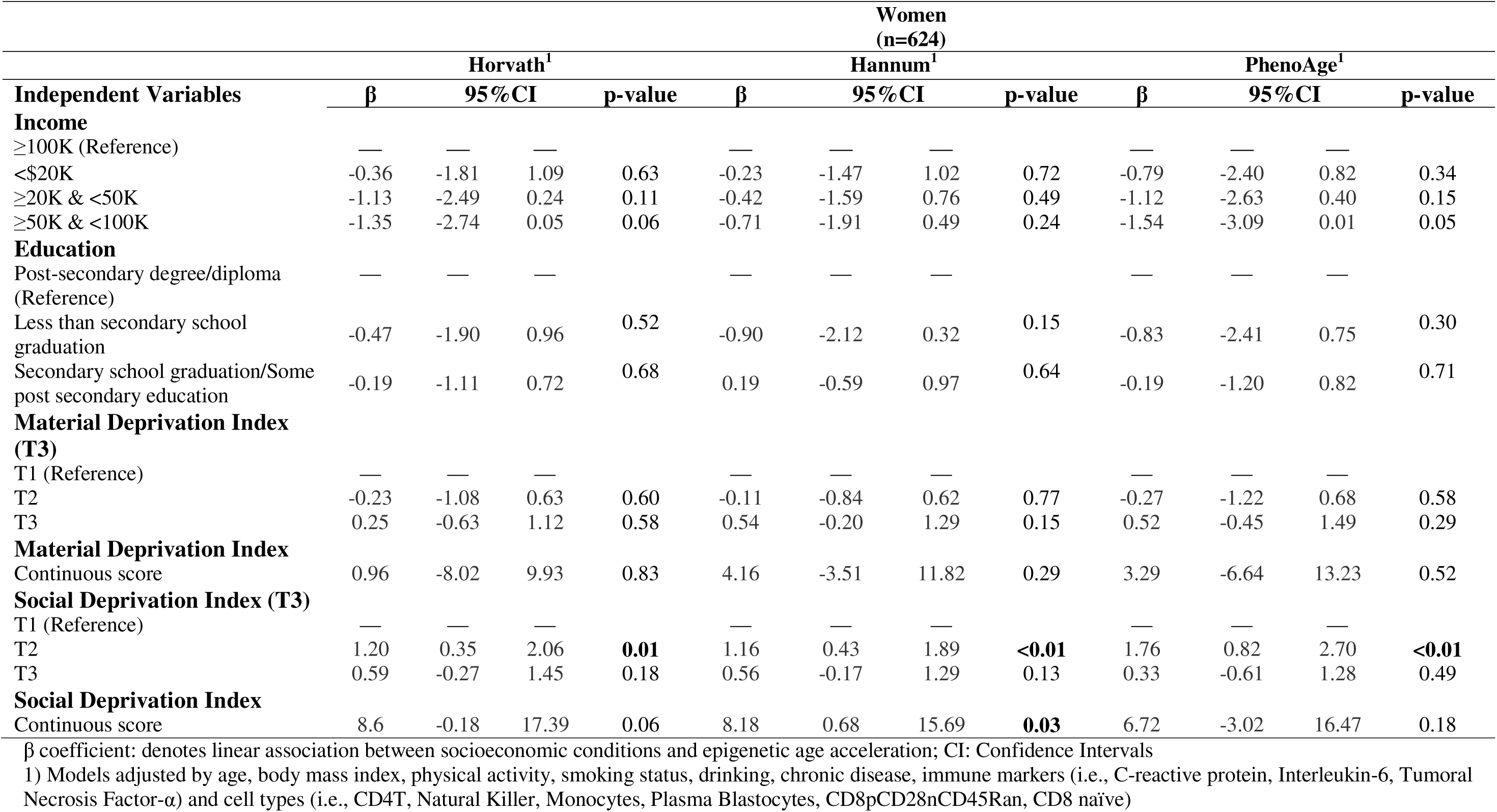
Association between DNA methylation age acceleration and socioeconomic conditions among female participants of the Canadian Longitudinal Study of Aging (CLSA)

**Supplemental Table 4:** Association between DNA methylation age acceleration and socioeconomic conditions among male participants of the Canadian Longitudinal Study of Aging (CLSA)

| Independent Variables | Men<br>(n=620) |  |  |  |  |  |  |  |  |  |  |  |
| --- | --- | --- | --- | --- | --- | --- | --- | --- | --- | --- | --- | --- |
|  | Horvath<sup>1</sup> |  |  |  | Hannum<sup>1</sup> |  |  |  | PhenoAge<sup>1</sup> |  |  |  |
| | $\beta$ | 95%CI | p-value | | $\beta$ | 95%CI | p-value | | $\beta$ | 95%CI | p-value | |
| **Income** |  |  |  |  |  |  |  |  |  |  |  |  |
| ≥100K (Reference) | — | — | — |  | — | — | — |  | — | — | — |  |
| <\$20K | -0.90 | -2.22 | 0.41 | 0.18 | -0.14 | -1.29 | 1.01 | 0.81 | 0.40 | -1.15 | 1.96 | 0.61 |
| ≥20K & <50K | -0.37 | -1.42 | 0.67 | 0.48 | -0.26 | -1.18 | 0.65 | 0.57 | -0.05 | -1.29 | 1.19 | 0.94 |
| ≥50K & <100K | -0.58 | -1.50 | 0.35 | 0.22 | -0.20 | -1.01 | 0.61 | 0.63 | 0.40 | -0.69 | 1.49 | 0.47 |
| **Education** |  |  |  |  |  |  |  |  |  |  |  |  |
| Post-secondary degree/diploma (Reference) | — | — | — |  | — | — | — |  | — | — | — |  |
| Less than secondary school graduation | 0.39 | -1.25 | 2.03 | 0.64 | 0.44 | -0.99 | 1.87 | 0.55 | 0.81 | -1.13 | 2.74 | 0.41 |
| Secondary school graduation/Some post secondary education | -0.18 | -1.13 | 0.78 | 0.72 | -0.55 | -1.38 | 0.28 | 0.20 | -0.38 | -1.51 | 0.74 | 0.50 |
| **Material Deprivation Index (T3)** |  |  |  |  |  |  |  |  |  |  |  |  |
| T1 (Reference) | — | — | — |  | — | — | — |  | — | — | — |  |
| T2 | -0.16 | -0.99 | 0.68 | 0.71 | -0.13 | -0.86 | 0.59 | 0.72 | -0.25 | -1.23 | 0.73 | 0.62 |
| T3 | -0.42 | -1.28 | 0.44 | 0.34 | -0.85 | -1.60 | -0.11 | **0.03** | -0.81 | -1.83 | 0.20 | 0.12 |
| **Material Deprivation Index Continuous score** |  |  |  |  |  |  |  |  |  |  |  |  |
|  | -1.37 | -10.14 | 7.4 | 0.76 | -7.73 | -15.35 | -0.12 | 0.05 | -5.43 | -15.76 | 4.91 | 0.30 |
| **Social Deprivation Index (T3)** |  |  |  |  |  |  |  |  |  |  |  |  |
| T1 (Reference) | — | — | — |  | — | — | — |  | — | — | — |  |
| T2 | 0.08 | -0.76 | 0.91 | 0.86 | 0.09 | -0.64 | 0.82 | 0.81 | -0.02 | -1.01 | 0.97 | 0.97 |
| T3 | 0.74 | -0.11 | 1.58 | 0.09 | 0.31 | -0.43 | 1.05 | 0.40 | 0.40 | -0.60 | 1.40 | 0.44 |
| **Social Deprivation Index Continuous score** |  |  |  |  |  |  |  |  |  |  |  |  |
|  | 8.63 | 0.37 | 16.9 | **0.04** | 2.75 | -4.47 | 9.97 | 0.46 | 6.23 | -3.54 | 16 | 0.21 |

$\beta$ coefficient: denotes linear association between socioeconomic conditions and epigenetic age acceleration; CI: Confidence Intervals
1) Models adjusted by age, body mass index, physical activity, smoking status, drinking, chronic disease, immune markers (i.e., C-reactive protein, Interleukin-6, Tumoral Necrosis Factor-α ) and cell types (i.e., CD4T, Natural Killer, Monocytes, Plasma Blastocytes, CD8pCD28nCD45Ran, CD8 naïve)

**Supplemental Figure 1:**
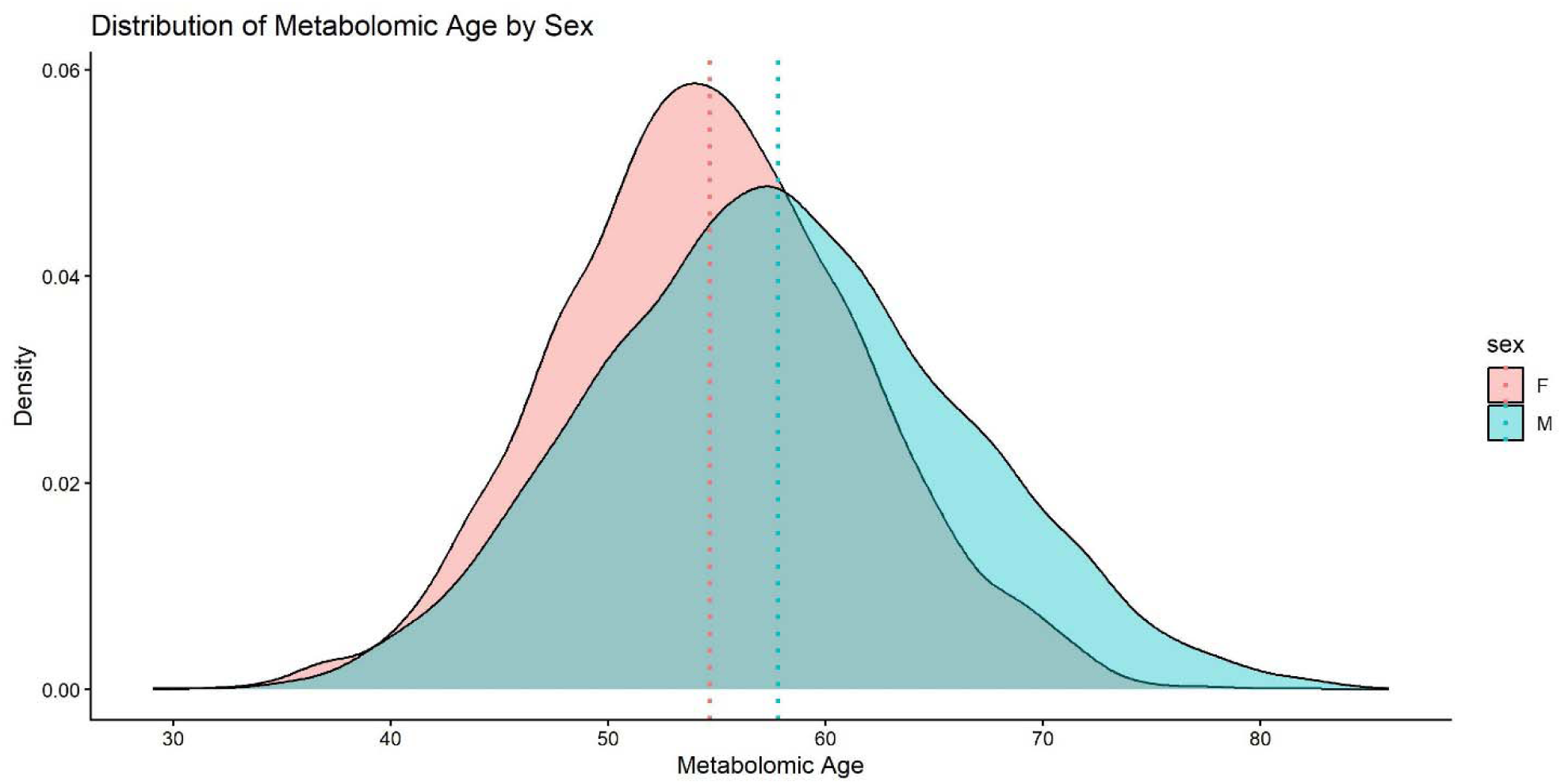
Density plot of metabolomic age by sex in the Canadian Longitudinal Study on Aging (CLSA, n=8,516). Dotted lines represent average metabolomic age for each sex (n=8,516).

**Supplemental Figure 2:**
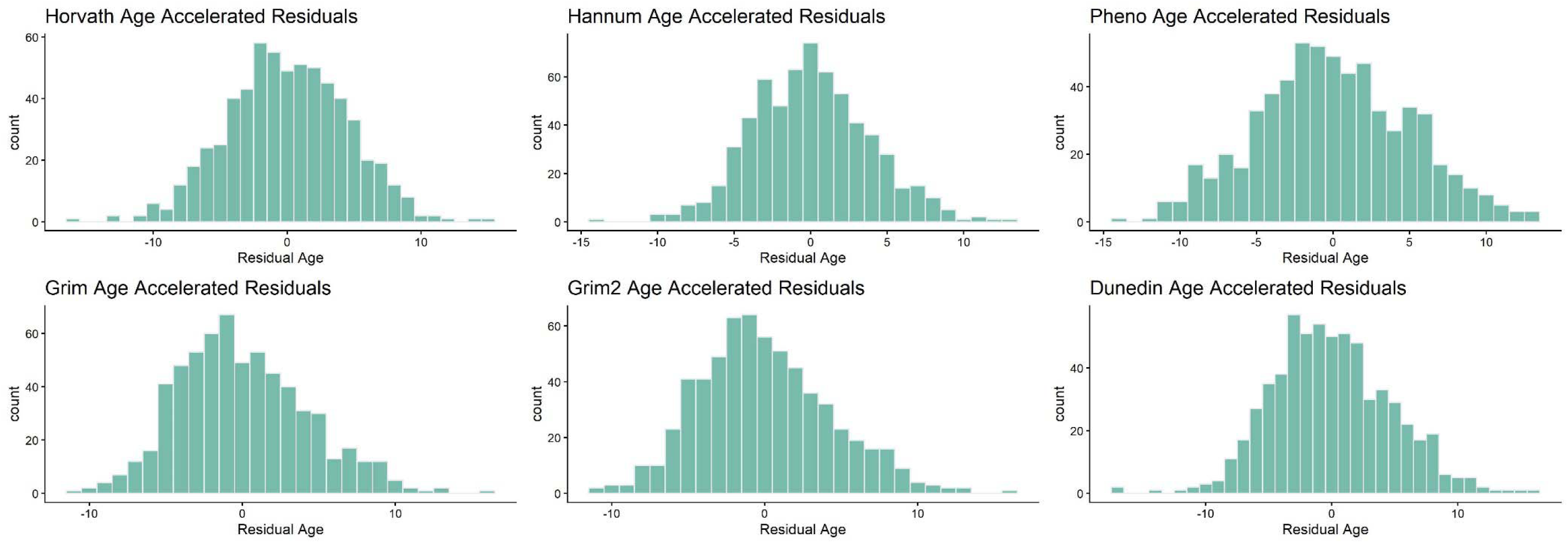
Histogram plots of epigenetic clocks age acceleration measures among females in the Canadian Longitudinal Study on Aging (CLSA, n=624).

**Supplemental Figure 3:**
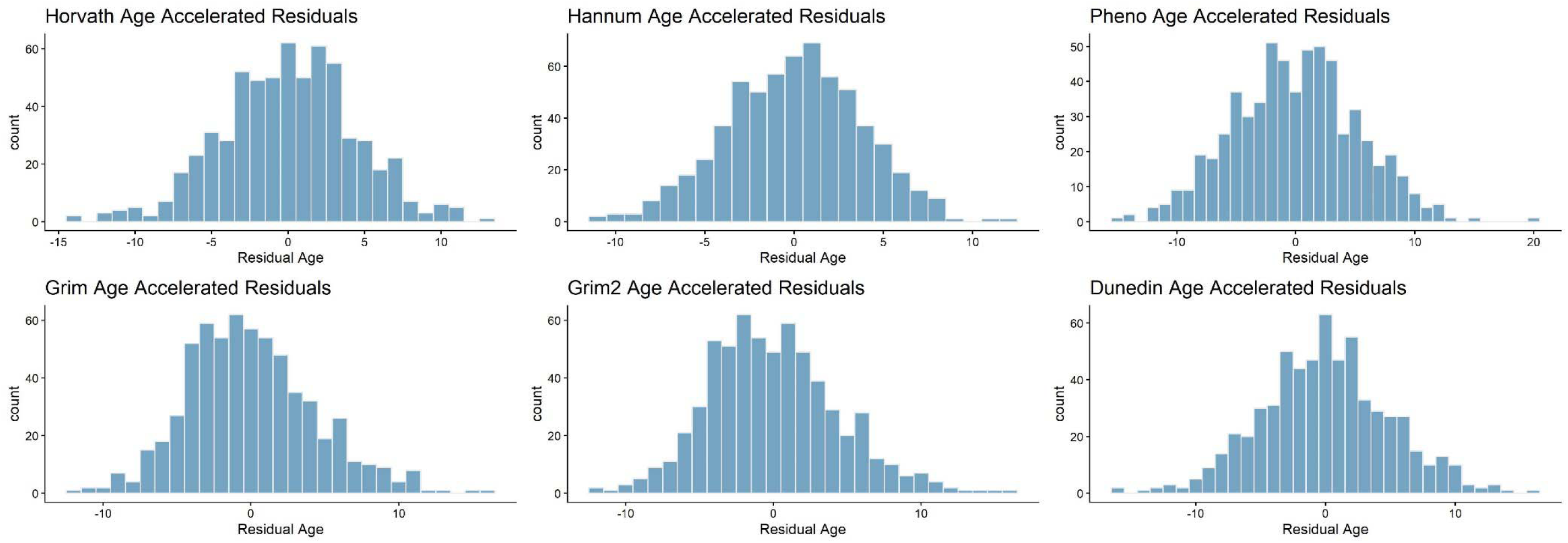
Histogram plots of epigenetic clocks age accelerated residuals among males in the Canadian Longitudinal Study on Aging (n=620).

## Notes

### Competing Interest Statement

The authors have declared no competing interest.

### Author Declarations

This study was approved by the IRB of Johns Hopkins University (IRB00441520)

